# Correlation of Axillary Nodal with Breast Response Following Neoadjuvant Chemotherapy in Early Breast Cancer

**DOI:** 10.64898/2026.10.14.26365456

**Authors:** A Agrawal, M Soliman, B Ahmed, Petros Barmpounakis, E Provenzano

**Affiliations:** Breast Surgery, Cambridge University Hospitals; School of Clinical Medicine, University of Cambridge; Surgical Oncology Department, Faculty of Medicine, Mansoura University, Egypt; Department of Pathology, Cambridge University Hospitals

**Keywords:** Breast cancer, axilla, lymph nodes, neoadjuvant chemotherapy, differential response

## Abstract

**Background:** Neoadjuvant chemotherapy (NACT) facilitates de-escalation of breast and axillary treatment in breast cancer, but pathological responses may differ between the primary tumour and axillary lymph nodes. Understanding differential response is crucial for de-escalation strategies.

**Methods:** This single university hospital retrospective study analysed operable breast cancers (cT1-cT3, cN0-cN3) treated with NACT (January 2013-November 2020). Two-tailed Fisher’s exact test or the Chi-squared test (for 2×2 comparisons) analysed concordance between breast and axillary pathological responses; Cox proportional hazards regression ascertained predictors of distant recurrence and survival; and logistic regression ascertained predictors of complete axillary response.

**Results:** Among 413 patients (cN0=229; cN+=184) with a median age of 51, pathological response concordance between the breast and axilla was 81% (both pCR, 22.2%; concurrent residual disease, 58.8%). The cN0 group had a significantly higher ypN0 rate in breast pCR versus non-pCR (97.1% vs 80.8%, p<0.001). cN+ had axillary pCR concordant with breast pCR, compared to only 16.5% with residual breast disease. Breast pCR was the most significant predictor of axillary pCR (OR=11.6, p<0.001). Axillary pCR was highest in ER-/HER2+(66.7%) and triple-negative(46.9%) subtypes. Residual axillary disease was independently associated with the worst breast cancer-specific (HR=7.13, p=0.006) and overall Survival (HR=3.32, p=0.029). Kaplan-Meier analyses showed a trend towards worse survival outcomes with axillary residual disease, even with a breast pCR.

**Conclusions:** In this limited series, breast pCR predicts axillary pCR in cN0 and cN+, particularly in HER2-positives and triple-negatives, supporting axillary de-escalation strategies in selected patients stratified by careful assessment of disease at the end of NACT.

## Introduction

Neoadjuvant chemotherapy (NACT) may downsize larger early breast cancer and facilitate breast conservation surgery (BCS)(Asselain et al., 2018). In more proliferative tumour subtypes (triple-negative and HER2-positive cancers), the magnitude of response to NACT provides essential prognostic information, with significantly improved survival associated with higher rates of pathological complete response (pCR). If, however, NACT is not an indication or results in a non-concentric or incomplete response, advancements in oncoplastic breast surgery techniques have recently permitted BCS (Ahmed et al., 2025).

However, beyond the primary breast, NACT may downstage the axilla in clinically node-positive (cN+) disease, enabling de-escalation of axillary surgery and reducing arm morbidity (Boughey et al., 2015). Emerging strategies such as tailored axillary surgery (TAS, removing biopsied/clipped nodes, sentinel nodes, and palpably suspicious nodes) or targeted axillary dissection (TAD, combining sentinel lymph node biopsy with clipped node excision) are reported to reduce false-negative rates to 2.4% (TAS) and 1.4% (TAD), respectively (Caudle et al., 2016). However, pathological responses in the breast and axilla may not be concordant. For instance, in a meta-analysis of 3,398 patients, persistent axillary disease was observed in 10% of those with breast pCR (Chehade et al., 2016). Hence, post-NACT but pre-operative axillary status is critical for subsequent axillary management.

We compared post-NACT responses in the primary breast tumour and the axillary lymph nodes, assessed predictors of axillary response, and focused on systemic oncological outcomes. This evidence will contribute to the emerging literature on response patterns and inform the selection of appropriate patients for both NACT and follow-on de-escalation strategies.

## Methods

### Study population and data collection

After obtaining institutional approval (PRN10914), we retrospectively analysed data of patients diagnosed between January 2013 and November 2020 with the following criteria:

Inclusions

- Over 18 years
- cT1-cT3 (operable breast cancer)
- cN0-cN3 disease
- Underwent NACT

Exclusions

- cT4 tumours
- Distant metastasis (Stage 4)
- Missing pathology data of either the breast or the axilla

Clinical, biological and histological information, including patient age, preoperative tumour size across imaging modalities, pre-treatment biological subtype, and final histological size and nodal status, was systematically extracted from Electronic Patient Records and compiled in a secure Microsoft™ Excel spreadsheet. Standard-of-care systemic therapy was administered to all patients guided by clinical parameters (Predict Online). For ER+/HER2-, this comprised Epirubicin and Cyclophosphamide with or without Fluorouracil, combined with Taxane-based chemotherapy (EC-T or FEC-T). For HER2+, chemotherapy was combined with anti-HER2-directed therapy (Trastuzumab); Pertuzumab was introduced in 2017. For TNBC, EC-T, FEC-T, or Platinum-based chemotherapy was used (before the advent of Immunotherapy). In the breast, complete response was defined as ypTis/ypT0, while in the axilla, it was defined as the absence of viable tumour cells within resected lymph nodes, including nodes showing fibrosis or acellular mucin pools consistent with treatment effect (ypN0).

### Aims & Objectives

A. Primary outcomes
  1. Pathological response types: complete (pCR), partial (pPR) and none (pNR) in the breast and axilla.
  2. Pathological response concordance rates between breast and axilla.
B. Secondary outcomes
  1. Tumour subtype analysis: stratification of NACT response.
  2. Predictors of complete axillary response.
  3. Clinicopathologic predictors of distant recurrence (DR), breast cancer-specific survival (BCSS) and overall survival (OS).
  4. Comparison of DR, BCSS and OS rates stratified by response in breast and axilla.

### Statistical analysis

Data were analysed using GraphPad Prism version 10.5.0. Descriptive statistics included counts and percentages for categorical data, and medians and interquartile ranges (IQRs) for continuous data. For two-by-two comparisons, two-tailed Fisher’s exact or Chi-squared tests were used. Univariable and multivariable Cox proportional hazards models were used to identify potential predictors of DR, BCSS, and OS. Univariable and multivariable logistic regression analyses were performed to identify predictors of complete axillary response. To assess concordance between breast and axillary pCR, McNemar’s test was used to assess the statistical significance of discordance. Kaplan–Meier survival curves were generated for DR, BCSS and OS, stratified by residual breast and axillary response, and included global log-rank p-values. Differences between groups were compared using pairwise log-rank tests with Benjamini-Hochberg FDR adjustment to control the false discovery rate. The statistical significance threshold was set at 5%. All survival and model analyses included cN+ patients, in addition to 20 cN0 patients with unexpectedly fibrosed nodes post-NACT (cN0/ypN0 with complete response) and 22 cN0 patients with unexpectedly positive SLNB (cN0/ypN+ with partial or no response).

## Results

Over 8 years, 413 patients with a median age of 51.2 years (IQR, 43.0-59.0) received NACT, and the median follow-up (from the post-NACT surgery date to the last data update; 25 March 2026) was 44.3 months (IQR, 28.1-59.5).

### Study population and tumour characteristics

**Table 1** presents patient demographics (n=413); the majority had cT2 tumours. 184 were node-positive (cN+), and 229 were node-negative (cN0). Standard reporting of progesterone receptor (PR) status is more recent (n=233). In HER2+ patients, Trastuzumab was used alone in 45% and in combination with Pertuzumab in 55%. In the TNBC cohort, platinum-based chemotherapy was used in 59%.

**Table 1:** Patient demographics and tumour characteristics.

|  |  |
| --- | --- |
| Median age in years (IQR) | 51.2 (43.0-59.0) |
| Median preoperative tumour size in mm (IQR) |  |
| Mammogram | 30.0 (22-42.75) (n=302) |
| Ultrasound | 25.0 (18.0-33.0) (n=368) |
| MRI | 38.0 (26-54.3) (n=356) |
| Clinical T staging | n=406 |
| cT1 | 15.8% (64/406) |
| cT2 | 67.2% (273/406) |
| cT3 | 17.0% (69/406) |
| Unknown | n=7 |
| Tumour grade | n=386 |
| 1 | 0.2% (1/386) |
| 2 | 30.8% (119/386) |
| 3 | 68.9% (266/386) |
| Unknown | n=27 |
| Focality | n=347 |
| Unifocal | 73.2% (254/347) |
| Multifocal | 26.8% (93/347) |
| Unknown | n=66 |
| Pathological T staging (ypT) | n= 412 |
| ypTis (residual DCIS only) | 9.5%(39/412) |
| ypT0 | 27.7% (114/412) |
| ypT1 | 35.7% (147/412) |
| ypT2 | 19.2% (79/412) |
| ypT3 | 7.3% (30/412) |
| ypT4 | 0.7% (3/412) |
| Unknown | n=1 |
| Tumour biology | n=413 |
| TNBC* | 31.2% (129 /413) |
| ER- HER2+ve | 10.4% (43 /413) |
| ER+ HER2+ve | 27.1% (112/413)<br>• ER+/PR+/HER2+ve<br>12.8% (53/413)<br>• ER+/PR-/HER2+ve<br>1.5% (6/413)<br>• Unknown=53 |
| ER+ HER2-ve | 31.2% (129/413) |
|  | ER+/**PR**+/**HER2**-ve<br>12.6% (52/413)<br>ER+/**PR**-/**HER2**-ve<br>4.8% (20/413)<br>Unknown=57 |
| Clinical Node stage (cN) | n=413 |
| Node positive (cN+) | 44.6% (184/413)<br>cN1=96.7% (178/184)<br>cN2=2.7% (5/184)<br>cN3=0.5% (1/184) |
| Node negative (cN0) | 55.4% (229/413) |
| Pathological Node staging (ypN) | n= 412 |
| ypN0 | 64.1% (264/412) |
| ypN0+ | 2.2% (9/412) |
| ypN1mi | 2.4% (10/412) |
| ypN1 | 19.2% (79/412) |
| ypN2 | 8.5% (35/412) |
| ypN3 | 3.6% (15/412) |
| Unknown | n=1 |
| Tumour subtype | n= 407 |
| NST | 89.4% (364/407) |
| ILC | 3.7% (15/407) |
| mixed | 3.2% (13/407) |
| Other | 3.7% (15/407) |
| unknown | n=6 |

NACT=Neo-adjuvant chemotherapy; ER= oestrogen receptor; PR= progesterone receptor; TNBC= triple negative breast cancer; NST= no special type; ILC= invasive lobular cancer
\*Includes 6 patients that were ER-/PR+/**HER2**-

### Pathological response rate and concordance rates between breast and axilla

**Table 2 summarises** pathological response rates and breast-axilla concordance. Breast pCR was higher in the cN0 group (cN0 45.4% vs. cN+ 26.2%) and nodal pCR [ypN0, 88.2% (201/228) vs. ypN+, 11.8% (27/228)]. Breast pCR was associated with ypN0 in 97.1% (100/103) of cN0 patients and with axillary complete response in 80.9% (38/47) of cN+ patients; conversely, residual breast disease was associated with ypN+ in 19.2% (24/125) of cN0 patients and with residual axillary disease in 84.1% (111/132) of cN+ patients. McNemar’s test indicated a significant imbalance in discordant pairs (p=0.009), demonstrating that breast and axillary pCR are not interchangeable measures of response. However, breast pCR was the most significant predictor of axillary pCR, with breast pCR being 11.6 times more likely to be associated with axillary pCR in multivariable analysis. Grade 3 was also a significant predictor of axillary pCR (OR=2.43), but tumour subtype and age were not (Supplementary Table 1). In the cN0 cohort, ypN0 was significantly more common with breast pCR than with breast pPR/pNR (97.1% vs. 80.8%, P <0.001).

**Table 2:** Pathological response rate in the breast and concordance between breast and axillary response (in cN0 and cN+ cohorts)

| Node negative group (cN0, n=229) |  | Node positive group (cN+, n=184) |  |
| --- | --- | --- | --- |
| Breast response | Axillary concordance | Breast response | Axillary concordance |
| **<u>Complete (pCR)</u>**<br>45.4% (104/229) | **<u>ypN0</u>**<br>97.1% (100/103)<br>**<u>ypN+</u>**<br>2.9% (3/103)<br>(ypN1mi = 2, ypN1 = 1) | **<u>Complete (pCR)</u>**<br>26.2% (48/183) | **<u>Complete</u>**<br>80.9% (38/47)<br>**<u>Partial</u>**<br>8.5% (4/47)<br>**<u>No response</u>** |
|  | **<u>Unknown/missed</u>**<br>n=1 |  | 10.6% (5/47)<br>**<u>Unknown/missed</u>**<br>n=1 |
| **Partial or No response (pPR or pNR)**<br>54.6% (125/229)* | **<u>ypN0</u>**<br>80.8% (101/125)<br>**<u>ypN+</u>**<br>19.2% (24/125)<br>(ypN0i+ = 3, ypN1mi = 7, ypN1=10, ypN2=3, ypN3=1) | **Partial or No response (pPR or pNR)\*\***<br>73.8% (135/183)<br>**<u>Unknown/missed</u>**<br>n=1 | **<u>Complete</u>**<br>15.9% (21/132)<br>**<u>Partial</u>**<br>13.6% (18/132)<br>**<u>No response</u>**<br>70.5% (93/132)<br>**<u>Unknown/missed</u>**<br>n=3 |

\* Only two patients (2/130) had no breast response
\*\*Only one patient (1/135) had no breast response (also had concomitant no axillary response)

### Tumour subtype analysis

**Figure 1 shows** response rates in the breast and axilla, stratified by tumour subtypes. **Supplementary Tables 1 and 2** provide detailed response rates. Breast pCR rates were higher in cN0 than in cN+ across all biological subtypes. In the cN+ cohort, nodal pCR rates exceeded breast pCR rates in all biological subtypes. In both the breast and axilla, ER-ve HER2+ve disease had the highest proportion of pCR, followed by TNBC. In cN+, ER-HER2+ tumours had 59.1% (13/22) breast pCR and 66.7% (14/21) axillary pCR, while TNBC had 39.4% (13/33) breast pCR and 46.9% (15/32) axillary pCR. The lowest complete response rates were observed in the ER+ve HER2-PR+ve subtype in both the breast (3%, 1/33) and axilla (11.8%, 4/34).

**Figure 1:**
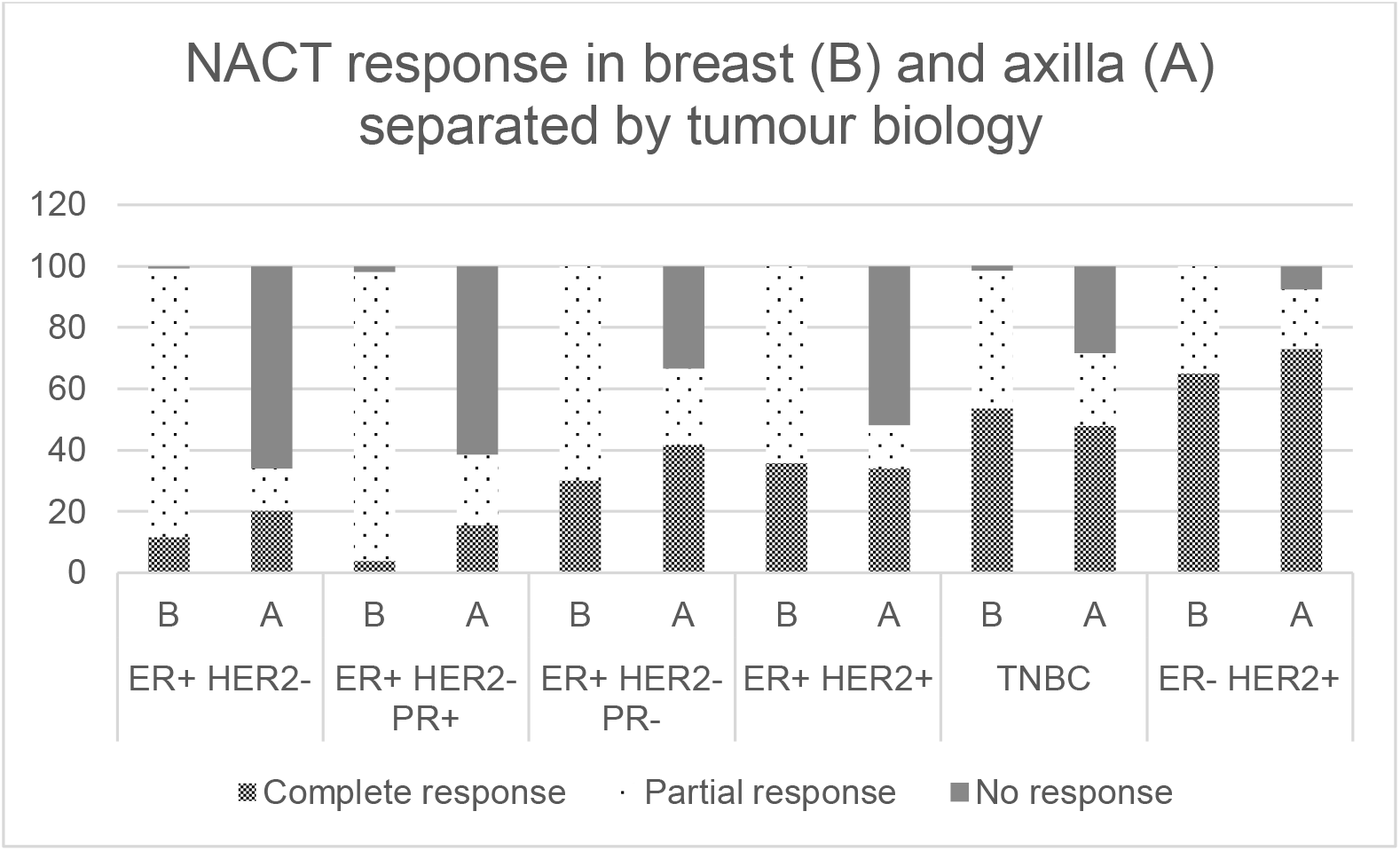
Bar-chart showing NACT response in breast (B) and axilla (A) in relation to breast pathological responses in all patients.

### Comparison of oncological outcomes

In our patient cohort, the overall mortality rate was 13.1% (54/413), and breast cancer-specific mortality was 9.7% (40/413). Local in-breast recurrence was 5.1% (3/59) in axillary pCR versus 9.1% (11/121) in partial or no response (p=0.55). **Table 3** presents hazard models for predictors of DR, BCSS, and OS. In the multivariable Cox model, patients with residual axillary disease had significantly higher breast cancer-related mortality (HR=7.13) and all-cause mortality (HR=3.32). TNBC and grade 3 tumours were significantly associated with a higher risk of distant recurrence. **Figure 2** shows Kaplan-Meier survival curves for DR, BCSS and OS, with a trend towards worse survival outcomes in those with axillary residual disease, even with a breast pCR. A pairwise log-rank test showed significantly worse outcomes for BCSS (p=0.026) and OS (p=0.043) with residual disease pattern, but not for DR. **Supplementary Table 3** outlines oncological outcomes according to breast and axillary responses and outcomes based on combined breast/axillary response in the cN+ cohort.

**Table 3:** Univariable and multivariable Cox proportional hazards models for distant recurrence, breast cancer-specific survival and overall survival.

|  | **Distant recurrence (DR) (n=148)** |  |
| --- | --- | --- |
| Variable | HR (95% CI) | P value |
| Residual axillary disease |  |  |
| - No (pCR) | - |  |
| - Yes | 2.03 (0.71-5.76) | 0.2 |
| Residual breast disease |  |  |
| - No (pCR) | - |  |
| - Yes | 2.29 (0.78-6.73) | 0.13 |
| Tumour subtype |  |  |
| - ER+ HER2- | - |  |
| - TNBC | 3.17 (1.17-8.61) | **0.023\*** |
| - HER+ ER- | 1.86 (0.31-11.1) | 0.5 |
| Tumour grade |  |  |
| - Grade 1 or 2 | - |  |
| - Grade 3 | 2.9 (1.23-6.83) | **0.015\*** |
| Age | 0.98 (0.95-1.02) | 0.3 |
|  | **Breast cancer-specific survival (BCSS) (n=201)** |  |
| Residual axillary disease |  |  |
| - No (pCR) | - |  |
| - Yes | 7.13 (1.75-29) | **0.006\*** |
| Residual breast disease |  |  |
| - No (pCR) | - |  |
| - Yes | 0.51 (0.16-1.67) | 0.3 |
| Tumour subtype |  |  |
| - ER+ HER2- | - |  |
| - TNBC | 1.95 (0.7-5.45) | 0.2 |
| - HER+ ER- | 0.53 (0.06-4.48) | 0.6 |
| Tumour grade |  |  |
| - Grade 1 or 2 | - |  |
| - Grade 3 | 1.97 (0.78-4.94) | 0.15 |
| Age | 1 (0.96-1.04) | 0.9 |
|  | **Overall Survival (OS) (n=201)** |  |
| Residual axillary disease |  |  |
| - No (pCR) | - |  |
| - Yes | 3.32 (1.13-9.75) | **0.029\*** |
| Residual breast disease |  |  |
| - No (pCR) | - |  |
| - Yes | 1.28 (0.45-3.65) | 0.6 |
| Tumour subtype |  |  |
| - ER+ HER2- | - |  |
| - TNBC | 1.72 (0.67-4.4) | 0.3 |
| - HER+ ER- | 1 (0.21-4.81) | >0.9 |
| Tumour grade |  |  |
| - Grade 1 or 2 | - |  |
| - Grade 3 | 2.29 (1.03-5.08) | **0.041\*** |
| Age | 0.99 (0.96-1.03) | 0.7 |

TNBC=Triple Negative Breast Cancer, CI = Confidence Interval, HR = Hazard Ratio

**Figure 2:**
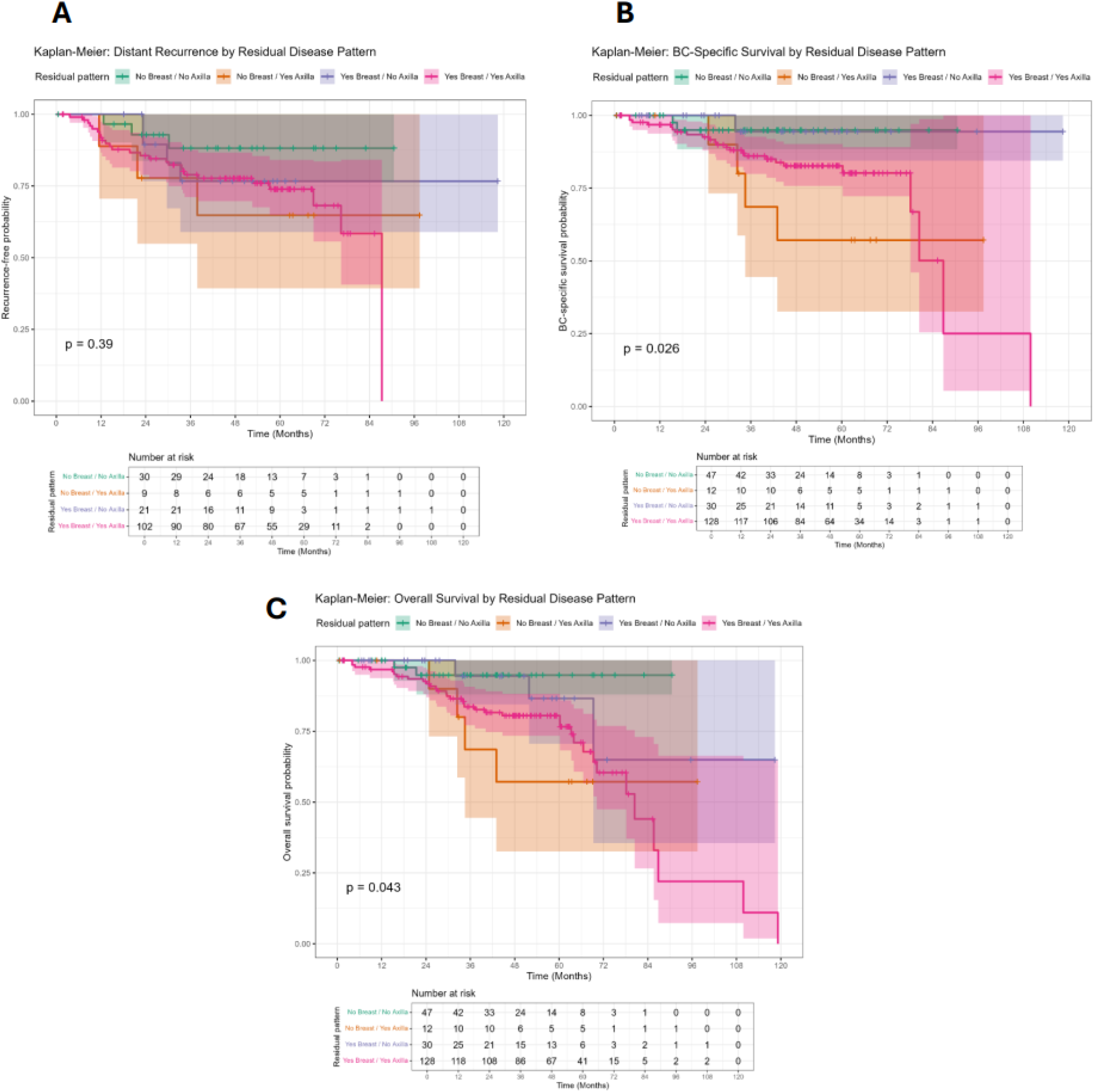
Kaplan–Meier survival curves for Distant Recurrence (A), Breast Cancer Specific Survival (B) and Overall Survival (C), stratified by breast and axillary response with global log-rank p-values (95% CI).

## Discussion

In our study, the overall breast pCR rate was 36.8% in the whole cohort (cN0 & cN+) and 26.2% in the cN+ cohort, consistent with the literature (breast pCR, 21% to 29%) (Hong et al., 2021; Lim et al., 2021), including a meta-analysis reporting a 22% rate (Cortazar and Geyer, 2015). These rates are expected to improve proportionally with better selection for NACT based on tumour biological subtypes predictive of favourable response (e.g., HER2+ and TNBC) (Cortazar and Geyer, 2015; Hong et al., 2021; Lim et al., 2021).

Breast pCR may predict negative axillary lymph node status in certain tumour subtypes, as concordant responses could support axillary-conservation strategies (Barron et al., 2018; Choi et al., 2019; Hong et al., 2021; Vugts et al., 2018). In cN0, breast pCR strongly correlates with ypN0 in both HER2+ and triple-negative subtypes, assuming the sentinel node biopsy false-negative rate is under 10% (Choi et al., 2019; Samiei et al., 2020).

### Concordance between breast and axilla and predictors of axillary response

The axillary pCR rate in our cN+ cohort was 33% (ACOSOG Z1071 rate, 41%) (Boughey et al., 2013). Pathological concordance (across all response types) between breast and axillary responses was 67.1%, with concordant pCR in 33.9% and concordant residual disease in 33.2% of patients. Morgan et al. reported a similar pathological concordance rate of 85.6% (pCR, 20.5%; residual, 65%) (Morgan et al., 2020). A large registry-based study from the American National Cancer Data Base (n=20,256) reported similar simultaneous pCR rates (19.2%) (Fayanju et al., 2018).

cN+ with breast pCR had a higher axillary pCR rate of 80.9% than those with residual breast disease (16%). Other studies have reported comparable axillary pCR rates of 74-89% (Choi et al., 2019; Hong et al., 2021; Morgan et al., 2020; Tadros et al., 2017). In multivariable analysis, breast pCR was the strongest predictor of axillary response (OR=11.6), followed by grade 3 tumours. There was a trend towards higher axillary response in ER-HER2+ (OR=2.88) and TNBC (OR=1.64).

Studies have reported a higher axillary pCR associated with breast pCR. In one study, the overall association was 4.5-fold (Samiei et al., 2020), whilst others were dominantly associated with ER-negative, HER2-positive, and grade 3 tumours (Choi et al., 2019; Hong et al., 2021; Youssef et al., 2021). This suggests that tumour biology is more important than tumour burden. Furthermore, Flores et al. reported that age >54, low tumour grade, and cN2/3 were significantly associated with higher odds of residual axillary disease despite breast pCR (Flores et al., 2023). Another study reported a lower rate of axillary pCR with higher T or N stage before NACT and low Ki67 expression (Hong et al., 2021). These data support axillary de-escalation, though emphasising careful consideration based on tumour biology.

In the cN0 group, only 2.9% (3/103) of breast pCR cases had unexpected ypN+ disease, compared with 19.2% (24/125) in those with residual breast disease (p<0.001). A meta-analysis of 9,609 cN0 patients with breast pCR reported an ypN+ rate of 0.2%. Stratified by subtype, ypN+ was lowest in ER+/HER2+ (0%), followed by ER-/HER2-(0.3%), ER-/HER2+ (0.6%), and ER+/HER2-(5.1%) (Ma et al., 2023). This supports evidence that breast pCR is highly predictive of axillary pCR in both clinically negative and positive axillae.

Predicting nodal involvement and response to NACT can be complex, relatively inaccurate, and dependent on several factors (Sivakanthan et al., 2024). Several traditional prediction models have been proposed for axillary response; however, their performance is often unsatisfactory (Kantor et al., 2018; Vila et al., 2016), and no accurate non-invasive technique exists (Schipper et al., 2015). Radiomics is an emerging interdisciplinary field that combines medical imaging with computer analysis to extract quantitative information from medical images and to build machine learning models. Some radiomics and deep learning models have demonstrated promising predictive performance for breast and axillary responses (Lin et al., 2024; Wang et al., 2024; Yu et al., 2024); however, their direct clinical application requires further work.

### Differential axillary response based on receptor profile

Not surprisingly, our study found the highest axillary pCR in ER-/HER2+ (66.7%) and TNBC (46.9%), and the lowest in ER+/HER2-(17.4%). Within ER+/HER2-cases, axillary pCR was slightly higher in PR-than in PR+ cases (36.4% vs 11.8%). PR positivity has previously been shown to increase the odds of nodal residual disease (OR⍰=⍰1.66, 95% CI⍰=⍰1.03–2.67, p⍰=⍰0.036) (Hong et al., 2021). Our results were comparable to a pooled meta-analysis of 57,531 patients, which reported axillary pCR rates of: ER-/HER2+ (60%), TNBC (48%), ER+/HER2+ (45%), and ER+/HER2-(18%) (Samiei et al., 2021). Other studies have also shown that, among those who achieved breast pCR, axillary pCR was highest in HER2+ and TNBC. In one study, breast pCR was associated with axillary pCR in 78-89% of HER2+ and 68-86% of TNBC, compared with 50-67% of ER+/HER2− cancers (Barron et al., 2018; Hong et al., 2021).

### Oncological outcomes with differential breast/axillary responses

This study demonstrates that concurrent breast and axillary pCR, and axillary pCR alone, were associated with superior BCSS and OS compared with breast pCR alone and residual breast and axillary disease. Among all factors, residual axillary disease was the most significant predictor of poorer survival outcomes (HR=7.13 for BCSS; HR=3.32 for OS). Similarly, a pooled analysis of 11,955 patients (Cortazar et al., 2014) shows that concurrent breast and axillary pCR (ypT0/is ypN0) improved event-free and overall survival compared with breast pCR alone, particularly in TNBC and ER-/HER2+ cancers. This underscores the significant impact of axillary downstaging on adjuvant treatment de-escalation and survival. Nodal status is the key prognostic marker, and pCR is a surrogate for survival; thus, nodal response should be considered alongside the likelihood of in-breast response. The recently published NSABP B-51 trial provides evidence that adding adjuvant regional nodal irradiation to ypN0 post-NACT does not improve invasive breast cancer recurrence or breast cancer-specific survival (Mamounas et al., 2025). A scoping review, including the NSABP B-51 and other studies, likewise concluded that there was no benefit from post-mastectomy or regional radiotherapy in cases with cN+/ypN0 (Cavalcante et al., 2025).

### Future applications and study limitations

Our study adds to the evidence that predicting post-NACT nodal status may not only provide prognostic information but also inform decisions on axillary management and adjuvant therapy. For instance, node-positive HER2+ or TNBC disease with breast pCR is more likely to show concordant axillary pCR and may be more suitable for axillary de-escalation, such as TAD/TAS, with lower odds of later conversion to completion axillary clearance. Furthermore, we observed a very low nodal positivity rate among cN0 patients with breast pCR, suggesting that significant de-escalation, or even complete omission, of axillary treatment in this subset could be considered in larger randomised prospective trials.

This study is limited by its single-centre retrospective design. Subgroup analysis of breast/axillary response concordance by molecular subtype could not be performed due to small subgroup sizes. Because axillary de-escalation surgeries, such as TAD/TAS, were not practised during the period of this cohort, this enabled detailed analysis of the axillary response (from axillary dissection pathology). To conclude, recommendations for NACT and subsequent de-escalation strategies must consider tumour biology, as an incomplete response can affect the surgical de-escalation pathway, both in the breast and especially in the axilla.

## Data Availability

All data produced in the present study are available upon reasonable request to the authors (institutional approval will need to be sought for such requests).

## Disclosures

### Data Access Statement

The data is observational from standard of care, and no new data was generated or available for peer review.

### Ethics Statement

This study is deemed a service evaluation/clinical audit by the NHS Health Research Authority Decision Tool (http://www.hra-decisiontools.org.uk/research/index.html). Individual patient consent was not needed.

## Acknowledgements

None

## Supplementary Tables and Figures

**Supplementary Table 1:** Univariable and multivariable logistic regression models predicting complete axillary response.

**Supplementary Table 2:** The complete, partial and no response rates in the breast and axilla, stratified by tumour subtypes in both cN0 and cN+ cohorts.

**Supplementary Table 3:** Oncological outcomes according to breast and axillary responses to NACT (3a) and outcomes based on combined breast/axillary response in the cN+ group (3b).

