## Supplemental Tables for "Correlation of Axillary Nodal with Breast Response Following Neoadjuvant Chemotherapy in Early Breast Cancer"

**Supplementary Table 1:** Univariable and multivariable logistic regression models predicting complete axillary response.

| **Variable** | **OR (CI)** | **P Value** |
| --- | --- | --- |
| Breast response   - Residual disease - Complete response (pCR) | -  11.6 (5.19-27.8) | **<0.001** |
| Tumour subtype   - ER+ HER2- - TNBC - HER+ ER- - HER2+ ER+ | -  1.64 (0.56-4.7)  2.88 (0.84-10.1)  0.99 (0.37-2.54) | 0.4  0.091  >0.9 |
| Tumour grade   - Grade 1 or 2 - Grade 3 | -  2.43 (1.07-5.85) | **0.038** |
| Age | 1.01 (0.97-1.04) | 0.7 |

TNBC=Triple Negative Breast Cancer, CI = Confidence Interval, OR = Odds Ratio

**Supplementary Table 2:** The complete, partial and no response rates in the breast and axilla, stratified by tumour subtypes in both cN0 and cN+ cohorts.

| **Tumour Biology** | **Region (Breast/Axilla)** | **Complete Response % (n)** | **Partial Response % (n)** | **No Response % (n)** |
| --- | --- | --- | --- | --- |
| TNBC | Breast (cN0) | 58.3 (56/96) | 40.63 (39/96) | 1.0 (1/96) |
|  | Breast (cN+) | 39.4 (13/33) | 57.8 (19/33) | 3.0 (1/33) |
|  | Axilla (cN+) | 46.9 (15/32) | 12.5 (4/32) | 40.6 (13/32) |
| ER+ HER2+ | Breast (cN0) | 36.2 (25/69) | 63.4 (44/69) | 0.0 (0/69) |
|  | Breast (cN+) | 34.9 (15/43) | 65.1 (28/43) | 0.0 (0/43) |
|  | Axilla (cN+) | 36.6 (15/41) | 7.3 (3/41) | 56.1 (23/41) |
| ER- HER2+ | Breast (cN0) | 71.4 (15/21) | 28.6 (6/21) | 0.0 (0/21) |
|  | Breast (cN+) | 59.1 (13/22) | 40.9 (9/22) | 0.0 (0/22) |
|  | Axilla (cN+) | 66.7 (14/21) | 23.8 (5/21) | 9.5 (2/21) |
| ER+ HER2- | Breast (cN0) | 18.6 (8/43) | 79.1 (34/43) | 2.3 (1/43) |
|  | Breast (cN+) | 8.2 (7/85) | 91.8 (78/85) | 0 (0/85) |
|  | Axilla (cN+) | 17.4 (15/86) | 12.8 (11/86) | 69.8 (60/86) |
| ER+ HER2- **PR+** | Breast (cN0) | 5.6 (1/18) | 88.9 (16/18) | 5.6 (1/18) |
|  | Breast (cN+) | 3.0 (1/33) | 97.0 (32/33) | 0.0 (0/33) |
|  | Axilla (cN+) | 11.8 (4/34) | 20.6 (7/34) | 95.8 (23/34) |
| ER+ HER2- **PR-** | Breast (cN0) | 44.4 (4/9) | 55.6 (5/9) | 0.0 (0/9) |
|  | Breast (cN+) | 18.2 (2/11) | 81.8 (9/11) | 0.0 (0/11) |
|  | Axilla (cN+) | 36.4 (4/11) | 27.3 (3/11) | 36.4 (4/11) |

TNBC=Triple Negative Breast Cancer

**Supplementary Table 3:** Oncological outcomes according to breast and axillary responses to NACT **(3a)** and outcomes based on combined breast/axillary response in the cN+ group **(3b).**

**Table (3a)**

| **Outcome** | **Complete Responders (CR)** | **Partial or non-responders** |
| --- | --- | --- |
| **Breast Response to NACT (n=412)** | | |
| Local Recurrence | 3.3% (5/152) | 8% (21/260) |
| Regional Recurrence | 1.3% (2/152) | 1.9% (5/257) |
| Distant Recurrence | 4.6% (7/152) | 16.2% (42/260) |
| Breast Cancer-Related Deaths | 4.6% (7/152) | 12.7% (33/260) |
| **Axilla Response to NACT (n=180)** | | |
| Local Recurrence | 5.1% (3/59) | 9% (11/121) |
| Regional Recurrence | 1.7% (1/59) | 2.0% (2/98) |
| Distant Recurrence | 6.8% (4/59) | 20.6% (25/121) |
| Breast Cancer-Related Deaths | 1.7% (1/59) | 19% (23/121) |

**Table (3b)**

| **Breast response** | | | | |
| --- | --- | --- | --- | --- |
|  | **Complete** | | **Partial/No Response** | |
|  | **Axilla response (n=176)** | | | |
|  | **Complete** | **Partial/No Response** | **Complete** | **Partial/No Response** |
| Local Recurrence | 0.0% | 8.3% (1/12) | 10.0% (3/30) | 10.0% (13/130) |
| Regional Recurrence | 0.0% | 8.3% (1/12) | 3.3% (1/30) | 2.3% (3/130) |
| Distant Recurrence | 6.1% (3/49) | 25.0% (3/12) | 13.3% (4/30) | 20.8% (27/130) |
| Breast Cancer-Related Deaths | 4.1% (2/49) | 33.3% (4/12) | 3.3% (1/30) | 26.2% (34/130) |
